# Common abnormality of gray matter integrity in substance use disorder and obsessive-compulsive disorder: A comparative voxel-based meta-analysis

**DOI:** 10.1101/2020.09.03.20187229

**Authors:** Benjamin Klugah-Brown, Chenyang Jiang, Elijah Agoalikum, Xinqi Zhou, Liye Zou, Qian Yu, Benjamin Becker, Bharat Biswal

**Author notes:** Corresponding authors Bharat Biswal Benjamin Becker.

## Abstract

**Aim:** To determine robust transdiagnostic brain structural markers for compulsivity by capitalizing on the increasing number of case-control studies examining gray matter alterations in substance use disorders (SUD) and obsessive-compulsive disorder (OCD).

**Design:** Pre-registered voxel-based meta-analysis of grey matter volume (GMV) changes through seed-based d Mapping (SDM), follow-up functional, and network-level characterization of the identified transdiagnostic regions by means of co-activation and Granger Causality (GCA) analysis.

**Participants:** Literature search resulted in 31 original VBM studies comparing SUD (n = 1191, mean-age = 40.03, SD = 10.87) and 30 original studies comparing OCD (n = 1293, mean-age = 29.18, SD = 10.34) patients with healthy controls (SUD: n = 1585, mean-age = 42.63, SD = 14.27, OCD: n = 1374, mean-age = 28.97, SD = 9.96).

**Measurements:** Voxel-based meta-analysis within the individual disorders as well as conjunction analysis were employed to reveal common GMV alterations between SUDs and OCD. Meta-analytic coordinates and signed brain volumetric maps determining directed (reduced or increased) brain volumetric alterations between the disorder groups and controls served as the primary outcome. Meta-analytic results employed statistical significance thresholding (FWE< 0.05).

**Findings:** Separate meta-analysis demonstrated that SUD (cocaine, alcohol, and nicotine) as well as OCD patients exhibited widespread GMV reductions in frontocortical regions including prefrontal, cingulate, and insular regions. Conjunction analysis revealed that the left inferior frontal gyrus (IFG) consistently exhibited decreased GMV across all disorders. Functional characterization suggests that the IFG represents a core hub in the cognitive control network and exhibits bidirectional (Granger) causal interactions with the striatum. Only OCD showed increased GMV in the dorsal striatum with higher changes being associated with more severe OCD symptomatology.

**Conclusions:** Findings demonstrate robustly decreased GMV across the disorders in the left IFG, suggesting a transdiagnostic brain structural marker. The functional characterization as a key hub in the cognitive control network and casual interactions with the striatum suggest that deficits in inhibitory control mechanisms may promote compulsivity and loss of control that characterize both disorders.

## 1. Introduction

Obsessive-compulsive disorder (OCD) and substance use disorder (SUD) represent two neuropsychiatric disorders characterized by maladaptive and persistent repetitive behaviors. Typically, OCD involves either hidden or overt ritualistic acts to obtain relief, whereas SUD engages in the consumption of a substance for rewarding effects or relief of distress. Initially, these behaviors serve a specific goal such as relief from emotional, physical, or social distress or rewarding experience. However, during the transition into the pathological state of the disorders, the initial goal-directed behavior becomes progressively habitual and ultimately compulsive. Although compulsivity represents a transdiagnostic key symptom of both disorders and overarching models emphasize the contribution of Pavlovian and instrumental learning mechanism to the development of compulsive behavior in both disorders (Robbins, Gillan, Smith, de Wit, & Ersche, 2012; Robbins, Vaghi, & Banca, 2019) the transdiagnostically shared neurobiological mechanisms of the disorders remain to be systematically examined. Also, SUD and OCD often co-occur (Blom et al., 2011; Lochner et al., 2014; Mancebo, Grant, Pinto, Eisen, & Rasmussen, 2009; Ruscio, Stein, Chiu, & Kessler, 2010) and co-morbidity has been reported to be a potential source of inefficient treatment (Glazier, Calixte, Rothschild, & Pinto, 2013).

Accumulating evidence from different lines of research suggests shared brain dysregulations between the disorders, such that both disorders have been characterized by dysregulations in central glutamatergic (Blom et al., 2011; Gass & Olive, 2008; Pittenger, Bloch, & Williams, 2011) and dopaminergic signaling which have been associated with key symptoms of both disorders as well as the regulation of behavioral control, associative learning and compulsivity (Bari & Robbins, 2013; Bellini et al., 2018; Cools, 2008). Moreover, functional neuroimaging studies emphasize an important role of frontocortical circuits in compulsive behavior and accumulating evidence suggests that neurofunctional dysregulations in specific frontocortico-striatal pathways facilitate the development of compulsive symptoms in both disorders (Ó. F. Gonçalves et al., 2016; Milad & Rauch, 2012; Vollstädt-Klein et al., 2010; Zhou et al., 2019).

Despite evidence from previous meta-analyses suggesting robust brain structural alterations in both, SUD and OCD patients relative to control subjects, shared and separable brain structural alterations between the disorders have not been systematically determined. Previous overarching conceptualizations and neuroimaging studies point to some candidate brain systems that have been identified most consistently, particularly frontostriatal circuits and cortical regions such as the insula (Everitt & Robbins, 2005; Goldstein et al., 2009; Koob & Volkow, 2010). Additionally, quantitative and qualitative voxel-base morphometry (VBM) studies have suggested altered gray matter indices in SUDs (see cocaine; (Crunelle et al., 2014; Ide et al., 2014; Rando, Tuit, Hannestad, Guarnaccia, & Sinha, 2013), Cannabis; (Cousijn et al., 2012; Wetherill et al., 2015), Alcohol; (Xiao et al., 2015; Yang et al., 2016) and Nicotine; (Hanlon et al., 2016; Zubieta et al., 2001)) and OCD (Lázaro et al., 2011; Radua, Van Den Heuvel, Surguladze, & Mataix-Cols, 2010; Rotge et al., 2010; So et al., 2008a) however, the shared gray matter alterations between the disorders have not been determined. Therefore, the determination of shared structural alterations between the disorders may not only facilitate to ascertain the neurostructural basis of compulsivity but may additionally enable the development of clinical interventions, including the determination of promising targets for invasive or non-invasive brain modulation techniques.

Against this background, the present study aimed at determining shared and robust brain structural markers for the disorders by capitalizing on the increasing number of case-control studies examining gray matter alterations in SUD patients and OCD patients relative to healthy control subjects. To this end, we combined original studies from three prevalent substances abused (Alcohol, Cocaine, and Nicotine) which we individually analyzed in a first step. Next, we investigated the shared brain gray matter alterations between SUDs and OCD via voxel-based meta-analysis. This meta-analytic approach has the potential to address the inconsistencies and lack of replicability that often characterizes the original studies on brain structural alterations in psychiatric disorders (Button et al., 2013; Ioannidis, 2011). Based on previous results from functional imaging studies and overarching models of compulsivity we hypothesized that SUDs and OCD will exhibit shared GMV alterations in frontostriatal regions.

Additionally, we aimed to further characterize the identified common region both on the behavioral and network level. To this end, we employed Neurosynth to identify functional co-activation networks of the identified region and employed Granger causality analysis (GCA) to resting-state data from an independent sample of healthy controls to further map the causal relationship between the identified region and the striatum.

## 2. Methods

The present meta-analysis followed the Preferred Reporting Items for Systematic Reviews and Meta-Analyses (PRISMA) (Moher et al., 2014) (shown in **Figure 1**) and the principles of conducting coordinate-based meta-analysis (Müller et al., 2018). This study has been pre-registered on the OSF repository (Registration DOI: 10.17605/OSF.IO/7YG6J). In the initial step, we identified original studies examining brain structural alteration in SUD (Cocaine, Alcohol, and Nicotine) and OCD by means of MRI-based voxel-based morphometry (VBM). For the literature search, four databases (PubMed, Web of Science, Neurosynth, and Scopus) were utilized and original articles were identified based on relevant references in review studies. Titles and abstracts returned by the search results were examined for subsequent full-text screening and inclusion. Only English language studies reporting whole-brain results in terms of coordinates (3-dimensions (x, y, z) and in Talairach or Montreal Neurological Institute stereotactic space) and published between the year 2000 to 2020 were included. The screening process resulted in original peer-reviewed studies employing case-control designs in SUD and OCD, respectively. The following search terms were applied: “Cocaine” OR “Cocaine use disorder” OR “Alcohol” OR “Alcohol use disorder” OR “Nicotine” OR “Smoking” OR “Obsessive-compulsive disorder” AND (Morphometry OR Voxel-based OR voxelwise). Only articles with case-control designs reporting differences between the respective diagnostic group and healthy control subjects were included. Additional exclusion criteria were as follows:(1) articles reporting only region-of-interest (ROI) results, (2) articles with poly-drug users and samples with high comorbidities with psychiatric or somatic disorders (e.g. schizophrenia or HIV), (3) articles focusing on parental drug exposure, and (4) articles reporting results from the same dataset from previous studies, 5 studies including samples lower than N = 10 per disorder.

**Figure 1.**
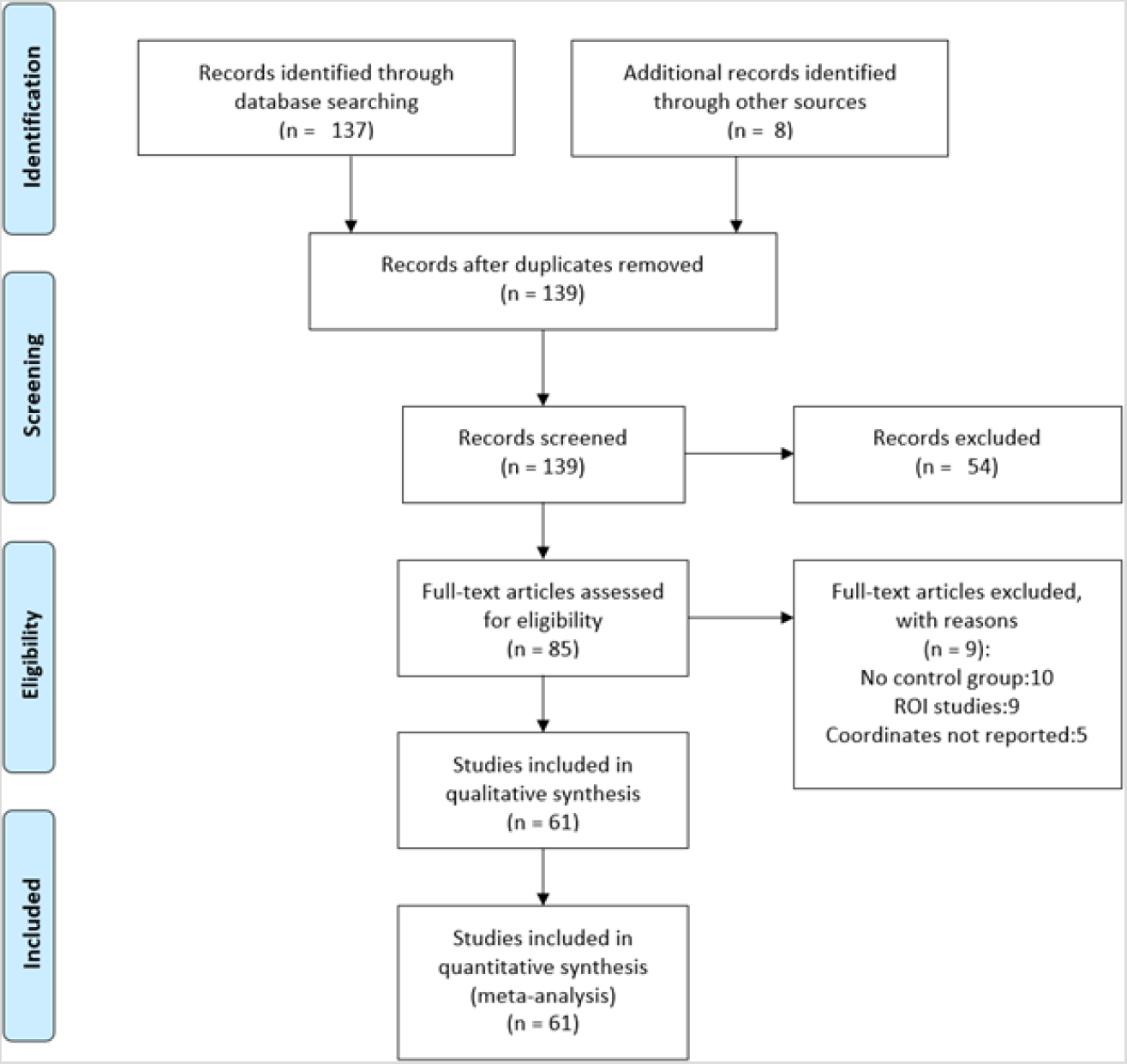
PRISMA. Selection of included studies

### 2.1 Meta-analytic approach

The meta-analysis of VBM studies was performed using SDM-PSI software version 2.1 (https://www.sdmproject.com/software/). Subsequent functional decoding of the identified regions was conducted via the Neurosynth database (https://neurosynth.org/). The analysis pipeline included the following steps: 1. Extraction of coordinates from peak clusters including their effective sizes (representing the gray matter differences between patients and controls) either in t-value, z-value or p-value (all values in z and p were converted to t-value using the statistical converter; https://www.sdmproject.com/utilities/?show=Statistics); 2. To account for differences between the reported coordinates and the standard space, we created MNI map of the GM for each study using the anisotropic Gaussian kernel with Full-width at half maximum set at 20mm and a voxel size of 2mm; 3. To account for potential effects of age in each original study mean of the samples was included as a covariate; and 4. Using the effective size maps and the sample sizes of each study variance maps were obtained. We next performed a voxel-wise computation to derive the mean maps by using the weighted mean study maps (obtained using the sample size, variances, and the between-study difference). In the analysis, we first computed the SDM for each disorder, and next a conjunction map was derived by computing the common regions to identify shared brain structural alterations between the four disorders. All meta-analytic results employed statistical significance testing by thresholding the derived maps by voxel-level uncorrected p< 0.001 and FWE< 0.05 (10 voxels) thresholds.

### 2.2 Sensitivity analysis

We performed whole-brain, voxel-based jackknife sensitivity analysis to determine the robustness of the results by setting a repetition value equal to the number of studies in each sample. The analysis in each of the groups was systematically repeated for 12, 9, 10, and 30 times, respectively, while discarding a single study each time. The number of repetitions equals the total number of studies in each sample, i.e. Alcohol(n = 12), Cocaine (n = 9), Nicotine (n = 10), and OCD (n = 30). This process was repeated until the last study was removed and placed back. The analysis was aimed at determining whether the observed findings are driven by single studies and thus testing the robustness of the group-level results.

### 2.3 Exploratory analyses of functional characterization: meta-analytic co-activation, causal connectivity and meta-regression

To functionally characterize regions exhibiting shared alterations across the disorders a co-activation analysis of the conjunction results across the diagnostic groups was performed. In the co-activation analysis, we used the inferior frontal gyrus (IFG) ROI co-ordinate to search for networks in the Neurosynth database, the result reflects regions extracted from a large database of previous studies that functionally co-activated with our ROI. Next, based on our *apriori* hypothesis of the importance of frontostriatal circuits in both disorders and compulsivity, we aimed to investigate causal relationships in the intrinsic interaction between the identified prefrontal (IFG) region and the striatum. Accordingly, resting-state fMRI data from n = 50 subjects (male;n = 28, mean age = 21.60, std = 2.01 and female; n = 22, mean age = 21,std = 2.24) all right-handed were included (for standardized resting-state data preprocessing see (Liu et al., 2019)) to examine the causal interaction of the left IFG with the ipsilateral striatum. We employed GCA (GCA derivation similar to (Benjamin Klugah-Brown et al., 2019)) to investigate the connectivity between the IFG and the targeted areas as it may allow us to explore the interactions between the identified region and the striatum on a causal level. For the technical details of GCA, please refer to the manuscript by Zang and colleagues (Zang, Yan, Dong, Huang, & Zang, 2012). Based on the meta-analytic determined overlapping GMV alterations between the SUDs and OCD, we defined the IFG as seeds (3mm-radius sphere) and striatum (left ventral and dorsal, seeds also known as targets) respectively. The striatal target-seeds were obtained from the human connectome atlas (https://atlas.brainnetome.org/bnatlas.html)and used 3mm radius ROIs for left striatum comprising of the dorsal and ventral striatum, respectively. Furthermore, voxel-wise, residual-based GCA evaluations were made on the mask of the grey matter using the REST toolbox (http://www.restfmri.net). The inflow (from the striatum to IFG), outflow (from IFG to striatum), and out-inflow (net flow) were also computed. The resultant inflow, outflow, and the net flow were further transformed to z-score to improve normality to facilitate the statistical analysis.

To further explore associations with parameters of symptom severity meta-regression analyses were performed. Given that the disorders are characterized by different core symptoms that are assessed via different measures we preformed meta-regression analyses for the duration of substance use for the SUD and Yale-Brown Obsessive Compulsive Scale YBOCs (severity score) for OCD. Note also that the variables are not the same throughout the four disorders, thus, apart from the usual confound (mean age and education), we used duration of substance use for the SUD and YBOCs (severity score) for OCD as predictors to examine associations, respectively. Spearman correlation with 95% confidence levels and thresholded at p< 0.05 were employed.

## 3. Results

Literature search performed according to our criteria resulted in a total of 31 original GM VBM studies in SUD (n = 1191, mean age = 40.03, SD = 10.87) and 31 OCD (n = 1293, mean age = 29.18, SD = 10.34) that compared brain structure via VBM to controls (SUD: n = 1585, mean age = 42.63, SD = 14.27, OCD: n = 1374, mean age = 28.97, SD = 9.96). The demographic characteristics of the samples from the included studies are presented in **Table 1**. There are no significant differences among the four groups (p< 0.05, F = 8.83). The breakdown of the 31 SUD study group comprising of three diagnostic categories is shown in **Table 2**.

**Table 1.**
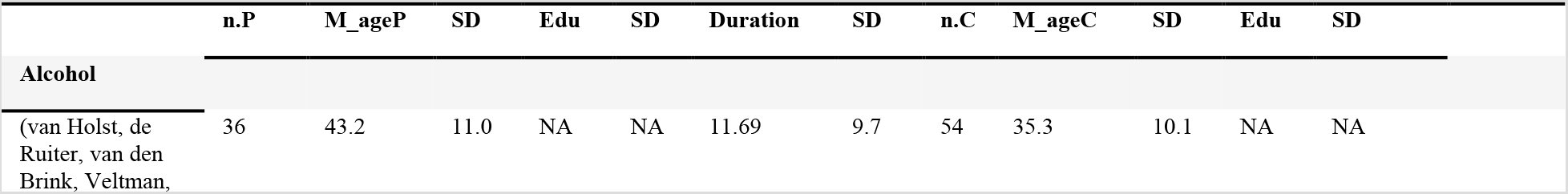
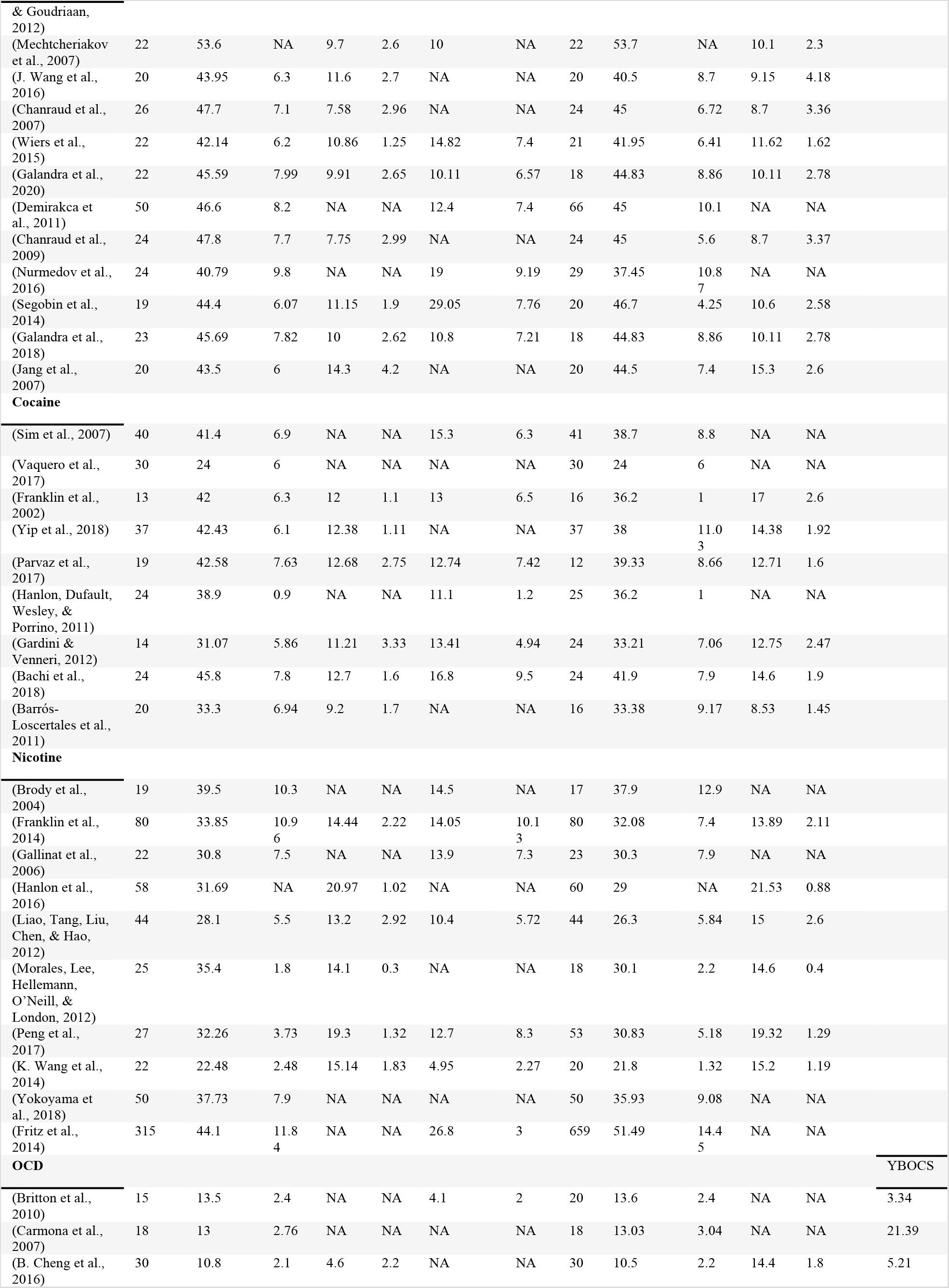
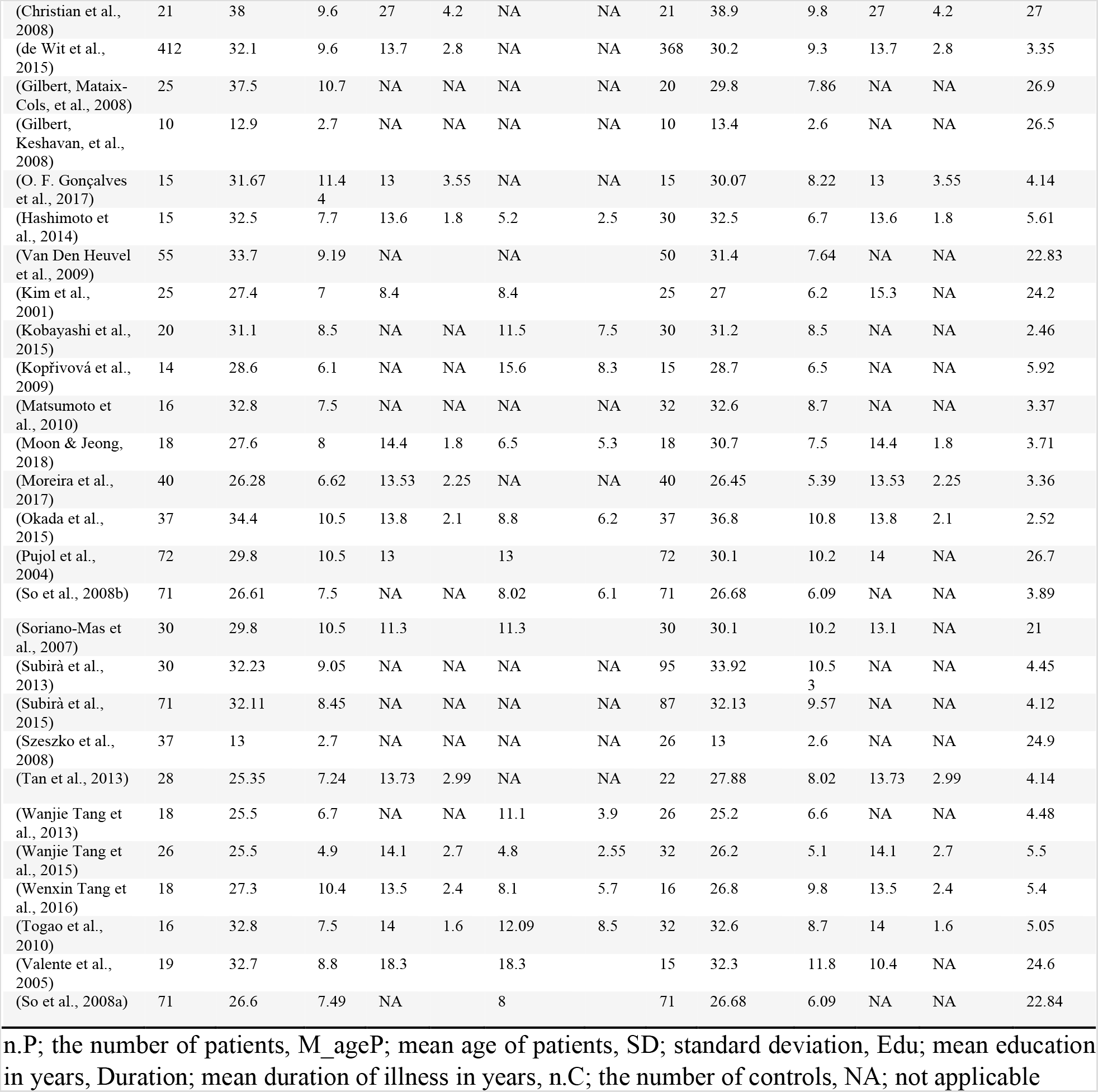
Demography of included studies

**Table 2.** Details of SUD category

| Category | SUD |  |  |  | Controls |  |  |  |
| --- | --- | --- | --- | --- | --- | --- | --- | --- |
|  | No. studies | Participants | Mean Age | Standard deviation |  | Participants | Mean Age | Standard deviation |
| Alcohol | 12 | 308 | 45.4685 | 8.2472 |  | 336 | 42.9538 | 9.6344 |
| Cocaine | 9 | 221 | 38.1661 | 9.2458 |  | 225 | 35.5804 | 9.2636 |
| Nicotine | 10 | 662 | 38.1255 | 11.5931 |  | 1024 | 44.0869 | 15.917 |

### 3.1 Main GMV Results

Results from the disorder-specific SDM-PSI meta-analysis are shown in **Figure 2**. After FWE correction was performed within each diagnostic category, reduced GMV for each diagnostic category as compared to healthy controls was mainly located in the bilateral anterior insula, and adjacent inferior frontal gyrus, dorsal anterior cingulate (dACC), and adjacent medial frontal gyrus (**Figures 2A-D**). All SUDs additionally exhibited reduced GMV of posterior insular regions (**Figures 2A-C)**, while nicotine use disorder exhibited reduced GM in the left, but not the right anterior insula (**Figure 1C**, z = 12). No regions with increased GMV were observed in any of the SUDs relative to healthy controls, whereas OCD showed increase GMV in the left striatum (putamen) (**Figure 4B**). An additional conjunction analysis in the three SUDs additionally revealed convergently decreased GMV in the left insula/IFG and the prefrontal cortex across the SUDs (results displayed in **supplementary Figure 1**). The conjunction analysis revealed that all four disorders exhibited convergently reduced GMV in the left inferior frontal gyrus (IFG) (**Figure 3A**). The detailed GMV changes in each disorder compared to healthy controls are presented in **Supplementary Table 1–4**. The results remained robust in the jackknife sensitivity analyses (**Supplementary Table 5**).

**Figure 2.**
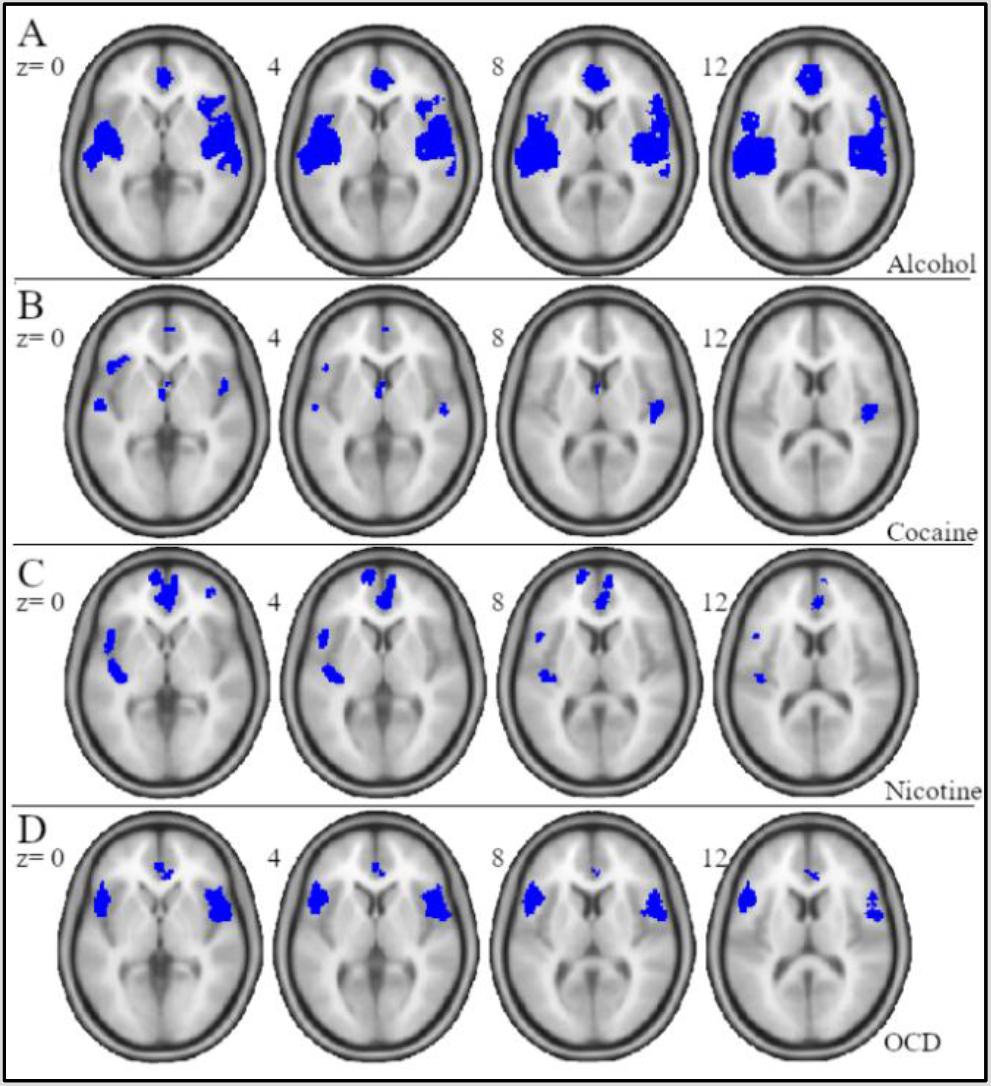
Figure 2Reduced GMV in each of the diagnostic group (patients < controls), corrected at FWE< 0.05

**Figure 3.**
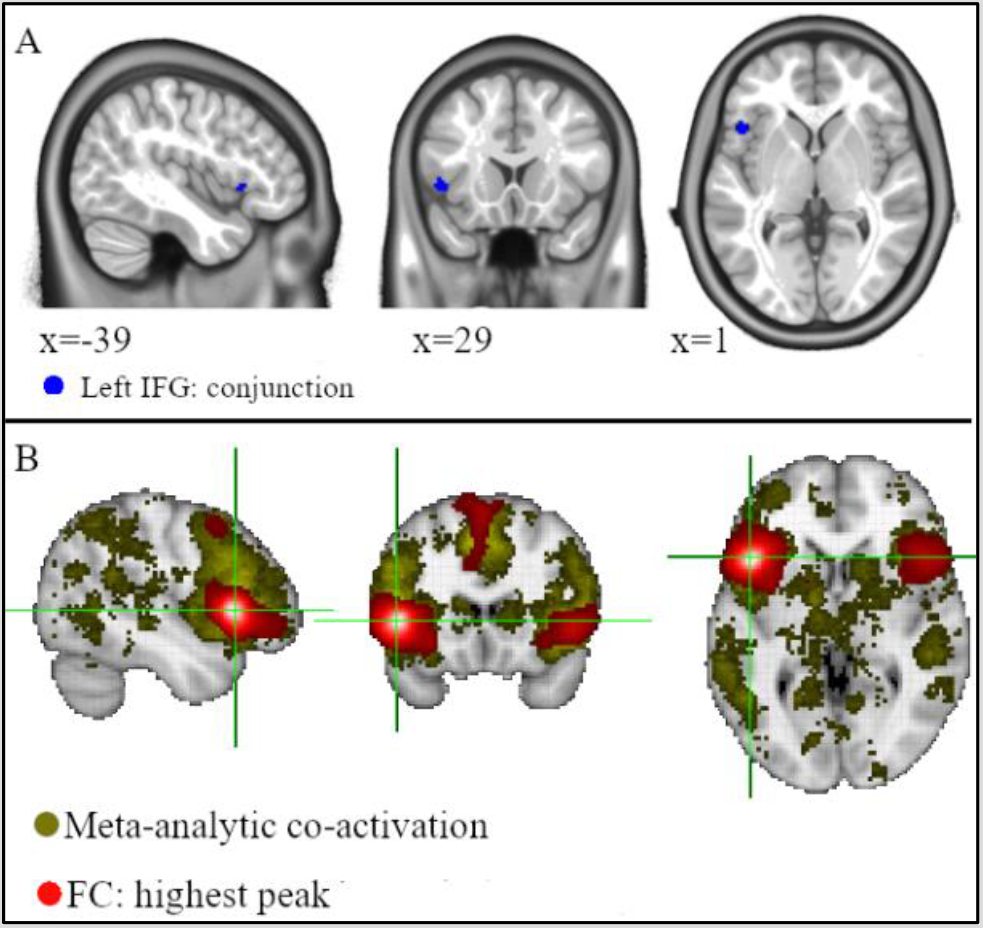
Conjunction and meta-analytic co-activation analysis. A) Reduced GMV overlap among the four groups (Patients < controls), corrected at FWE< 0.05. B) shows regions coactivated with the seed (IFG)

**Figure 4.**
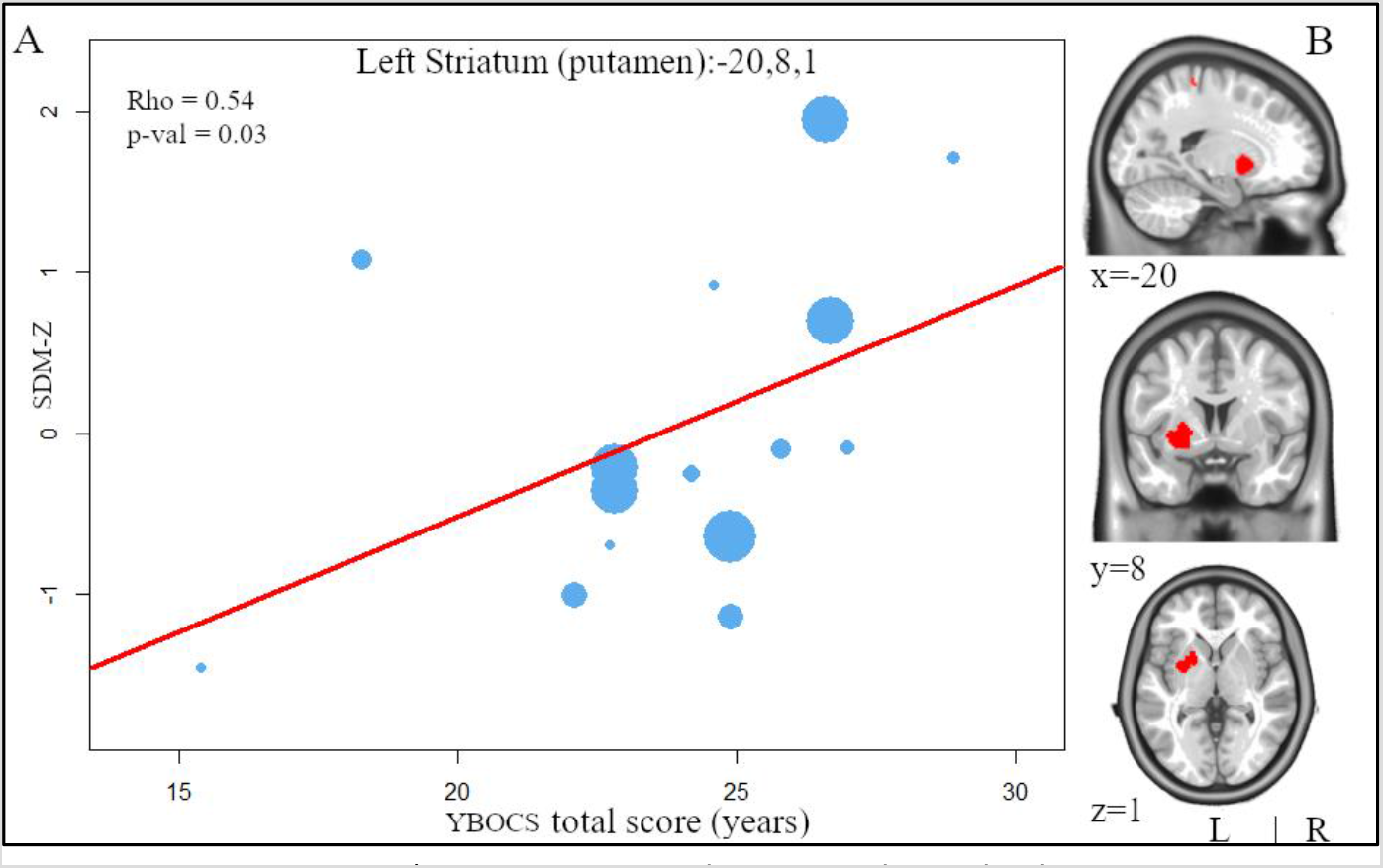
Meta-regression. A) Meta-regression showing a relationship between OCD severity (measured using mean Yale-Brown Obsessive-Compulsive Scale (YBOCS) scale) and GMV in left putamen. Each dot shows studies included in the regression; the different sizes symbolizes greater sample sizes. The meta-regression SDM-Z values indicates the proportion of studies that reported GM alterations close to the voxel. B) Increase GMV in OCD: L. Putamen, corrected at FWE< 0.05. Significant clusters were overlaid on an mni_icbm152 template for display purposes only

### 3.2 Exploratory analyses: meta-regression and neural decoding

To functionally characterize the region identified, a functional co-activation analysis was conducted. Coactivation analysis revealed that the identified IFG region primarily co-activated with broad regions of the frontal and parietal regions encompassing mainly the frontoparietal control network (**Figure 3B**).

The meta-regression revealed no significant associations (age, duration of substance use for SUD). However, in OCD, a significant association with symptom severity was found in the left dorsal striatum. Specifically, a higher OCD symptom as assessed by YBOCS scores were associated with increased left putamen GMV in the OCD patients (**Figure 4A**).

Findings from the GCA in the sample of healthy subjects revealed that causality was observed between the IFG and the striatum, specifically, an inflow pattern from left striatum regions to the IFG except for left dorsal putamen (dPu.L) (p< 0.05), **Figure 5A**. Generally, the pattern of bidirectional causal flow indicated a higher inflow from the target regions to the IFG (p< 0.05) with inflow/outflow between IFG and dCa.L showing the highest causality. In these results, we found that there was only positive NetFlow from the seed region to the left ventral putamen (vPu.L), signifying the causal influence from the IFG to this subregion of the striatum (**Figure 5B**).

**Figure 5.**
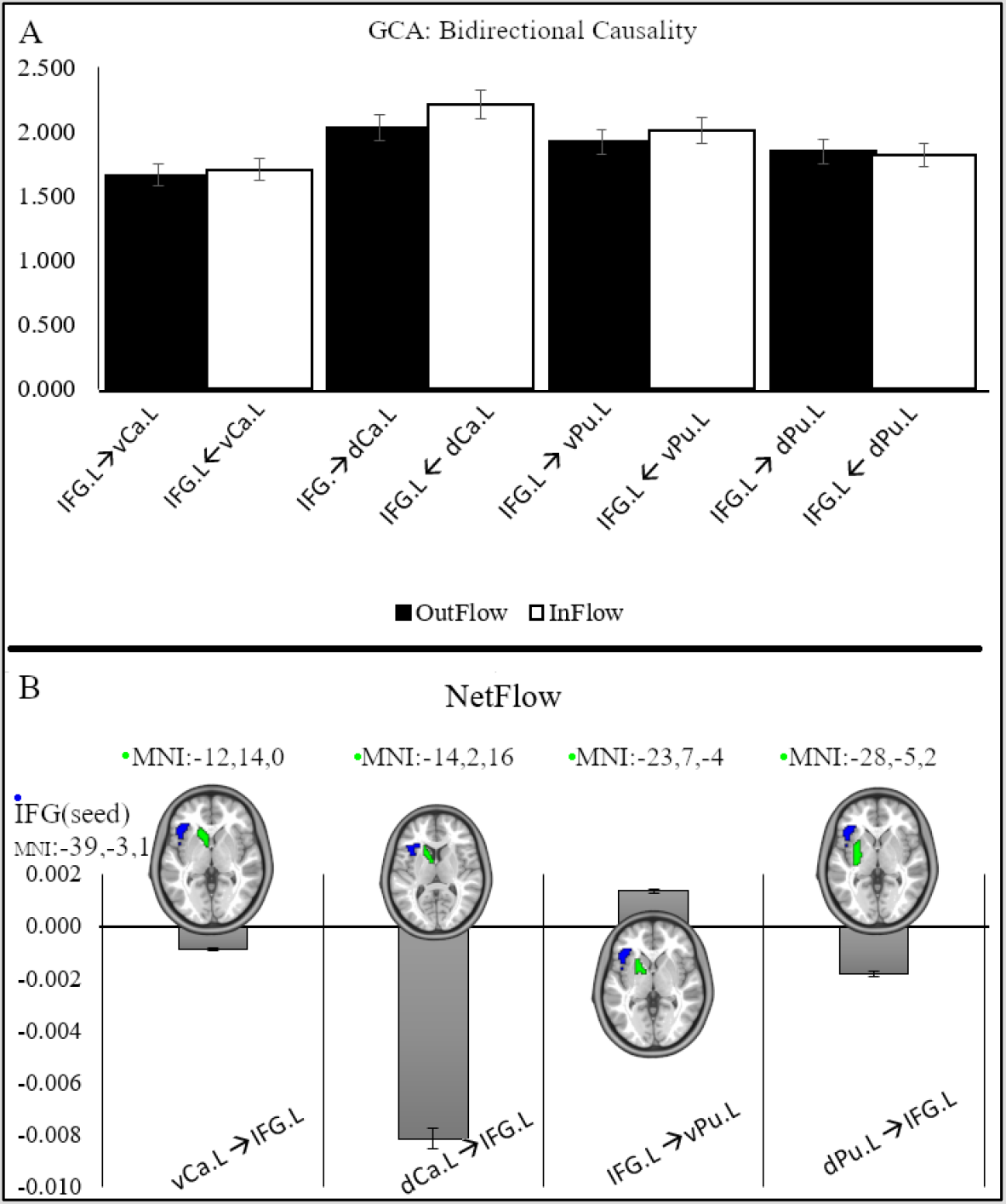
Causality between seed region and target ROIs. A) Each group of bars represent GCA residual computation from seed region IFG to selected striatal regions and vice versa. B) the overall causality between the IFG and the striatum regions. Seed region computed from the conjunction GMV of all diagnostic disorders’y-axis values are the z-scores computed from the residual GCA. Target ROI extracted from Brain connectome project atlas. vCa; ventral caudate, dCa; dorsal caudate; vPu; ventral putamen, dPu; dorsal putamen. The error bar shows the percentage standard error. MNI are the center location of the region

## 4. Discussion

The present meta-analytic study examined for the first time shared GMV alterations between SUDs and OCD. The disorder-specific voxel-wise meta-analysis revealed widespread medial frontal and insular GMV reductions within the SUD and OCD compared to controls and the conjunction meta-analysis revealed that the disorders are transdiagnostically characterized by reduced GMV in the left IFG. Subsequent exploratory analysis that aimed at functionally characterizing the identified region revealed that the left IFG functionally co-activated with a broad network including bilateral parietal and frontal regions, suggesting that this region represents a core node in the frontoparietal control networks. Based on the consistently reported role of frontostriatal circuits in SUD as well as OCD the intrinsic causal influence of the identified IFG region over the striatum was examined in an independent dataset of healthy subjects and revealed that the causal influence propagated from the left striatum to the left IFG, whereas the IFG exerted causal influence over the ventral putamen. Finally, we found that only OCD exhibited increased GMV, specifically in the left dorsal striatum (putamen) which positively correlated with OCD symptom severity (as assessed by the YBOCs) in OCD. The findings generally remained robust when subjected to jackknife sensitivity analyses. Together, the meta-analytic approach allowed us to determine common GMV alterations between SUD and OCD which may underly the shared symptoms on the behavioral level, specifically compulsive behavior and loss of behavioral control which characterizes both disorders.

### 4.1 Implications - SUD

The frontocortical GMV decreases in the SUD groups broadly resemble findings from the previous meta-and mega-analysis in subjects with SUD (e.g. (Ersche, Williams, Robbins, & Bullmore, 2013; Mackey et al., 2019; Yang et al., 2016)). Across the SUDs examined, the insula showed marked decreases in the GMV. The insula alteration was functionally shown in our previous study suggesting common neurofunctional alterations in this region across SUDs (B. Klugah-Brown et al., 2020) as well as between SUDs and OCD (Klugah-brown et al., 2020). The insula has increasingly been noted as addiction relevant region, probably via its important role in interoceptive processing, decision making, and/or risky behavior which may promote substance abuse despite being aware of the negative consequences (B. Klugah-Brown et al., 2020; Naqvi & Bechara, 2009). Structural deficit and/or functional alteration of the insula has been repeatedly described and associated with an increased relapse risk (Paulus, Tapert, & Schulteis, 2009). Moreover, the SUDs were characterized by medial frontal GM decreases, a region involved in decision making, self-awareness and regulatory control, such that deficits in this region have been shown to promote dysregulated reinforcement (Bechara, Tranel, & Damasio, 2009; Mackey et al., 2016). Decreased GMV in this network thus may neurally accompany the relationship between maladaptive decision, self-awareness and deficient regulatory control characterizing addictive disorders.

### 4.2 Implications – OCD

In OCD, widespread regions of the bilateral insula and focused regions in the IFG as well as dorsal anterior cingulate exhibited decreased GMV. On the functional level, the frontocortical and cingulate regions have consistently been found to be disrupted in OCD patients relative to controls in a number of fMRI studies examining resting-state (Y. Cheng et al., 2013; de Vries et al., 2019; Swedo et al., 1989; Yun et al., 2017) and task-based (Friedman et al., 2017; Maltby, Tolin, Worhunsky, O’Keefe, & Kiehl, 2005; Marsh et al., 2014; Yücel et al., 2007) neural activation. Our meta-analysis in OCD patients determined a decreased GMV in a similar network and previous studies have linked the identified network to cognitive control and inhibitory control mechanisms (Chamberlain, Blackwell, Fineberg, Robbins, & Sahakian, 2005; Yücel et al., 2007). Additionally, our meta-regression revealed an association between higher symptom scores in OCD patients and GMV increases of the dorsal striatum (putamen), a region that has been previously reported in functional neuroimaging studies in OCD (Baxter et al., 1996; Rapoport & Wise, 1988; Saxena, Brody, Schwartz, & Baxter, 1998).

### 4.3 Implications – common decreases in IFG GMV

The principal aim of the present meta-analysis was to determine shared GMV alterations between the disorders to facilitate the identification of brain structural commonalities which may underpin compulsivity, thus representing a key transdiagnostic symptom across SUDs and OCD. The corresponding conjunction analysis revealed that the left IFG exhibited shared volumetric decreases across the disorders. The left insular and adjacent IFG is known to play an important role in regulatory top-down control, particularly response inhibition (Devito et al., 2013), a neurocognitive function that has been found to be impaired in both disorders and may promote the development of compulsive behavior (Chamberlain et al., 2005; Zilverstand, Huang, Alia-Klein, & Goldstein, 2018). Moreover, the subsequent exploratory analysis that aimed at functionally characterizing this region suggests that the identified IFG region co-activates with widespread regions in the parietal and frontal cortex that highly resembles the front-parietal cognitive control network critically engaged in executive functions and behavioral control (Chen et al., 2018; Dixon et al., 2018; Fiske & Holmboe, 2019; Gürsel, Avram, Sorg, Brandl, & Koch, 2018; Reineberg, Gustavson, Benca, Banich, & Friedman, 2018; Stern, Fitzgerald, Welsh, Abelson, & Taylor, 2012). Based on previous conceptualizations and studies proposing a critical engagement of the frontostriatal circuits in both, SUD and OCD we employed GCA to examine the causal relationship between the identified IFG region and the striatum. Results confirmed a causal information flow between the two structures, specifically left striatal subregions causally influenced the IFG whereas the IFG controlled the ventral putamen. With respect to SUD, both the ventral striatal and the dorsal striatum have been engaged in maladaptive reward processing and the development of compulsive behavior (Andrews et al., 2011; Patel et al., 2013). Thus, the causal relationships and the main conjunction meta-analytic result support the suggestion that compulsivity and altered response to reward and/or punishment may be linked via these pathways that have been involved in both addiction and OCD. Interestingly previous studies have demonstrated shared alterations in this circuit between OCD and behavioral addiction (pathological gambling (Scherrer, Xian, Slutske, Eisen, & Potenza, 2015)), suggesting a transdiagnostic functional dysregulation in these pathways and some previous deep-brain stimulation studies have shown that compulsive behavior in OCD and addiction can be significantly attenuated through decreasing frontostriatal connectivity (De Ridder, Vanneste, Kovacs, Sunaert, & Dom, 2011; Dunlop et al., 2016; Figee et al., 2013; Kravitz et al., 2015; Valencia-Alfonso et al., 2012).

Summarizing, shared GMV loss in a region of the IFG which represents a principal node in the cognitive control network and critically interacts with the striatum may characterize addictive disorders and OCD and may underly the compulsive behavior exhibited by both disorders. The findings, therefore, resonate with previous conceptualizations proposing shared neurobiological alterations between the disorders that may promote the development of transdiagnostic symptoms of compulsivity.

It is noteworthy that, despite the important insights that the meta-analytic approach may have allowed, some limitations hindered the full examination of the topic. Firstly, only a limited number of VBM have been conducted in SUD resulting in a comparably low number of studies for the separate SUDs. However, across the separate SUDs convergent changes in the insula and prefrontal cortex were observed suggesting substance-independent GM alterations in SUD and pooling the data for the comparison with OCD increased the statistical power. Nevertheless, findings in the individual SUDs need to be interpreted cautiously. Secondly, since the disorders require different diagnostic symptom assessments, a meta-regression with severity measures could not be conducted across all disorders. Also, studies in each category did not report consistent measures (some studies did not report severity/duration and other measures). We, therefore, encourage researchers to report these measures as it reflects the core relationships between altered regions and symptoms.

## 5 Conclusion

We capitalized on previous case-control VBM studies in three prevalent SUDs and OCD with the aim to determine shared brain structural alterations across the disorders. The left IFG exhibited decreased GMV across all disorders suggesting a transdiagnostic marker that may underly the key symptomatic feature of compulsivity that characterizes the disorders. The IFG plays an important role in inhibitory control and our findings indicate that this region functionally interacts with both, the cognitive control network and the striatum, suggesting that this region plays a key role in the interaction between frontal regulatory control functions and habitual and reward-driven behavior. The findings emphasize that the symptomatological overlap may be rooted in common brain alterations and may open a new venue towards transdiagnostic treatment approaches that target brain alterations that promote compulsive behavior.

## Data Availability

All data associated with this work shall be provided upon request

## Acknowledgment

This work was supported by the National Key Research and Development Program of China (Grant No. 2018YFA0701400), National Natural Science Foundation of China (NSFC, No 91632117), and Science, Innovation and Technology Department of the Sichuan Province (2018JY0001).

